# Lack of Epistatic Interaction of *SNCA* with *APOE* in Synucleinopathies

**DOI:** 10.1101/2024.08.12.24311821

**Authors:** Prabhjyot Saini, Eric Yu, Mehrdad A. Estiar, Lynne Krohn, Kheireddin Mufti, Uladzislau Rudakou, Jennifer A. Ruskey, Farnaz Asayesh, Sandra B. Laurent, Dan Spiegelman, Isabelle Arnulf, Jacques Y. Montplaisir, Jean-François Gagnon, Alex Desautels, Yves Dauvilliers, Gian Luigi Gigli, Mariarosaria Valente, Francesco Janes, Andrea Bernardini, Karel Sonka, David Kemlink, Wolfgang Oertel, Karri Kaivola, Annette Janzen, Giuseppe Plazzi, Elena Antelmi, Francesco Biscarini, Michela Figorilli, Monica Puligheddu, Brit Mollenhauer, Claudia Trenkwalder, Friederike Sixel-Döring, Valérie Cochen De Cock, Christelle Charley Monaca, Anna Heidbreder, Luigi Ferini-Strambi, Femke Dijkstra, Mineke Viaene, Beatriz Abril, Bradley F. Boeve, Ronald B. Postuma, Guy A. Rouleau, Victoria Anselmi, Abubaker Ibrahim, Ambra Stefani, Birgit Högl, Michele T.M. Hu, Sonja W. Scholz, Ziv Gan-Or

## Abstract

Two recent studies suggested that the *APOE* ε4 haplotype was associated with increased α-synuclein pathology in cell and mouse models. Genetic variants in the *SNCA* region have strong association with Parkinson’s disease (PD), Dementia with Lewy Bodies (DLB), and idiopathic REM Sleep Behavior Disorder (iRBD), while *APOE* is a genetic risk determinant for only DLB. To determine if genetic-level interactions between *SNCA* and *APOE* exists that can explain the protein-level association, we investigated the genotypic interaction of *APOE* and *SNCA* in cohorts of PD, DLB, and iRBD. We analyzed genome-wide association study (GWAS) data from 5,229 PD patients and 5,480 controls, 2,610 DLB patients and 1,920 controls, and 1,055 iRBD patients and 3,667 controls. We used logistic regression interaction models across all 3 cohorts independently between the 1) top GWAS signals of *SNCA* SNPs and *APOE* haplotypes, 2) SNP x SNP and 3-way SNP interaction across the entire coding region plus 200kb flanking each gene. No significant interactions were found to be associated with any of the synucleinopathies after correction for multiple testing. Our results do not support a role for genetic interactions between *APOE* and *SNCA* across PD, DLB, and iRBD. Since the tested genetic variants affect the expression and function of these proteins, it is likely that any interactions between them does not affect the risk of PD, DLB and iRBD.

## 1. Introduction

Alpha-Synucleinopathy is an umbrella term to describe several neurodegenerative diseases that have a common defining pathological feature, characterized by neuronal or glial inclusions of aggregated alpha-synuclein, known as Lewy bodies, Lewy neurites or glial cytoplasmatic inclusions in the brain^1^. Disorders that are collectively referred to as alpha-synucleinopathies include Parkinson’s disease (PD), Dementia with Lewy Bodies (DLB) and multiple system atrophy (MSA). Furthermore, there is an increased presence of alpha-Synucleinopathy in the prodromal condition idiopathic/isolated REM sleep behavior disorder (iRBD), which can convert to either PD, DLB or MSA in more than 80% of cases^2^.

Alpha-synuclein is encoded by the *SNCA* gene, and genetic variants in the *SNCA* locus are associated with PD, DLB and iRBD risk in genome-wide association studies (GWASs)^3-8^. Specifically, some variants of *SNCA* are strongly associated with PD (rs356182 and rs2870004), while others are associated with DLB (rs7681440 and rs7680557)^7,9^ and iRBD (rs2870004)^10^. The top *SNCA* association in PD is independent and different than the top associations in DLB^7^ and iRBD^8^, raising the hypothesis that there could be differential effects of *SNCA* variants on the expression of alpha-synuclein in different brain regions^8^.

Overlapping neuropathologic features associated with Alzheimer’s disease (AD) are seen in the brains of many patients with PD^11^ and dementia including amyloid plaques composed of amyloid-beta (Aβ) plaques and neurofibrillary tangles containing the tau protein and may contribute to clinical features of disease^12,13^.

Coding variants in apolipoprotein E (*APOE*) produce 3 common alleles, ε2, ε3, and ε4. The *ε4* allele of *APOE* regulates lipid metabolism and cholesterol transport and known to be a major genetic risk determinant for sporadic, late-onset Alzheimer’s Disease^14^ and Lewy Body Dementia^7,9^. Dose effects by allele have demonstrated a 3.7-fold risk of developing AD while homozygosity increases the risk by up to 12-fold^7^. From numerous GWASs, A*POE* does not alter the risk for PD, yet the ε4 allele has been described as a potential risk factor for cognitive decline and development of dementia in PD patients^15,16^.

Two studies in mice demonstrated that the APOE*ε4 genotype was associated with increased alpha-synuclein pathology, independent of the amyloid β deposition^17,18^. These two studies emphasize a potential molecular mechanism of APOE*ε4 on α-Synuclein protein aggregation. However, beyond A53T, they have not evaluated disease specific variants of *SNCA* that have functional molecular consequences (*e*.*g*. E46K)^19^. Analysis of these variants could be insightful in understanding molecular association between *SNCA* variants and APOE*ε4. Furthermore, while the *SNCA* locus is associated with all synucleinopathies, *APOE* is a genetic risk factor for DLB only. *SNCA* and *APOE* variants may affect the expression / function of the proteins encoded by them^20^. Therefore, if a true interaction exists at the protein level as suggested by the studies mentioned above, then there plausibly should be evidence of some association at the genetic level.

Genetic interactions refer to a combination of two or more genetic variants whose phenotypic contribution is amplified by their co-occurrence^21^. To determine if genetic-level interactions between *SNCA* and *APOE* exist that can explain the protein-level association as described, we investigated the genotypic interaction of *APOE* and *SNCA* in three disease cohorts of PD, DLB and iRBD patients and controls, with a total of 8,855 patients and 11,067 controls.

## 2. Methods

### Patient population

For PD, we used the International Parkinson’s Disease Genomics Consortium (IPDGC) data set that contained 10,709 subjects with 5,229 cases and 5,480 controls. For DLB, we used the most recent DLB GWAS dataset which included 4,530 subjects with 2,610 cases and 1,920 controls. The iRBD cohort was composed of 4,742 individuals, including 1,055 cases and 3,667 controls with 1,968 controls from NGRC, (dbGAP: phs000196.v2.p1) and 790 controls from NIND (dbGAP: phs000089) added from external studies of Parkinson’s patients in addition to the controls collected for the iRBD cohort. PD was diagnosed using UK Brain Bank criteria or Movement Disorders Society (MDS) criteria^4^. DLB patients were diagnosed with pathologically definite or clinically probable disease according to consensus criteria^22^. iRBD was diagnosed according to the International Classification of Sleep Disorders (2^nd^ or 3^rd^ Edition)^23,24^. Informed consent and ethics approval was obtained from the appropriate institutional review boards at participating institutions as described in the original studies.

### Genetic Analysis

We generated genotype calls of two *APOE* SNPs, rs429358 and rs7412 to determine the *APOE* haplotype status of each sample. The combination of genotypes for rs429358 (C/T) and rs7412 (C/T) defines the three *APOE* haplotypes: epsilon 2 (ε2), epsilon 3 (ε3), and epsilon 4 (ε4). These three haplotypes can produce six genotypes, ε2/ε2, ε2/ε3, ε2/ε4, ε3/ε3, ε3/ε4, and ε4/ε4, whose frequency amongst the cohorts is detailed in Table 2.

Cohorts were collected, quality controlled, genotyped and filtered on individual and variant-level as previously described for PD^4^, iRBD^8^, and DLB^7^. *APOE* variants were analyzed for the 200kb region flanking both sides on chromosome 19: 44,705,791 - 45,109,393 and for *SNCA* on chromosome 4: 89,500,345 - 90,038,324. For the iRBD cohort, additional cohorts of control subjects added from NIND and NGRC were included. The DLB genotypic data was converted from GrCh38 to GrCh37 using Liftover^17^. Because external controls were added to the iRBD cohort; the cases and control genotypes were filtered for minor allele frequency (MAF) > 0.01 to reduce imputation errors and imputed using Michigan Imputation Server and the Haplotype Reference Consortium^18^ r1.1 2016 reference panel (GRCh37/hg19). Only imputed genotypes with an *R*^2^ > 0.30 were kept for analysis. Additionally, prior to analysis further quality control for each cohort included removing duplicate samples, missing data including covariates; SNPs were filtered based on variant missingness (< 0.05), genomic relatedness (> 0.125), disparate missingness between cases and controls (*p* > 1E-04), missingness by haplotype (*p* > 1E-04), deviation from Hardy-Weinberg equilibrium (*p* > 1E-04), minor allele frequency (MAF) > 0.01, and LD pruned with r^2^ at >0.5 with a 50kb window using plink 1.9 ^19^.

### Statistical Analysis

Descriptive measures of mean, standard deviations, frequencies, and percentages were used to summarize the data. SNP and haplotype interaction were analyzed using logistic regression controlling for age, sex and ancestry using the first five principal components. Epistasis model was defined as

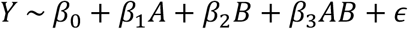

where *β*_0_ represents the intercept, *β*_*n*_ represents the coefficients for each SNP, and A and B represent allele dosage of each SNP and AB represents the interaction. The test for interaction was based on the coefficient *β*_3_ and P-value<0.05 as significant when testing interaction between top SNPs, and Bonferroni threshold applied for SNP x SNP interactions. Detecting gene-gene interactions in complex diseases can be accomplished using a variety of epistasis-focused tools or packages in existing software packages. Model-based Multifactor Dimensionality Reduction (MB-MDR) was implemented as a final screen strategy to detect any significant SNP x SNP and SNP x SNP x SNP interactions using the open-source MB-MDR v 4.4.1 software, as MB-MDR merges multi-locus genotypes exhibiting some significant evidence of High or Low risk, based on association testing into a new lower-order dimension. A new association test is subsequently performed per marker pair/triplet, by adopting a permutation-based strategy that corrects for multiple testing (over all marker pairs/triplets) and adequately controls family-wise error rate at α=5%^20^.

## 3. Results

To explore possible interactions between *APOE* and *SNCA* in cases and controls, we first focused primarily on the top *SNCA* SNPs associated with PD, DLB, and RDB (Table 1) and the *APOE* haplotypes. We performed a SNP-haplotype logistic regression interaction controlling for age, sex and ancestry using the first five principal components. No significant interactions were found to be statistically significant (Table 1).

**Table 1:**
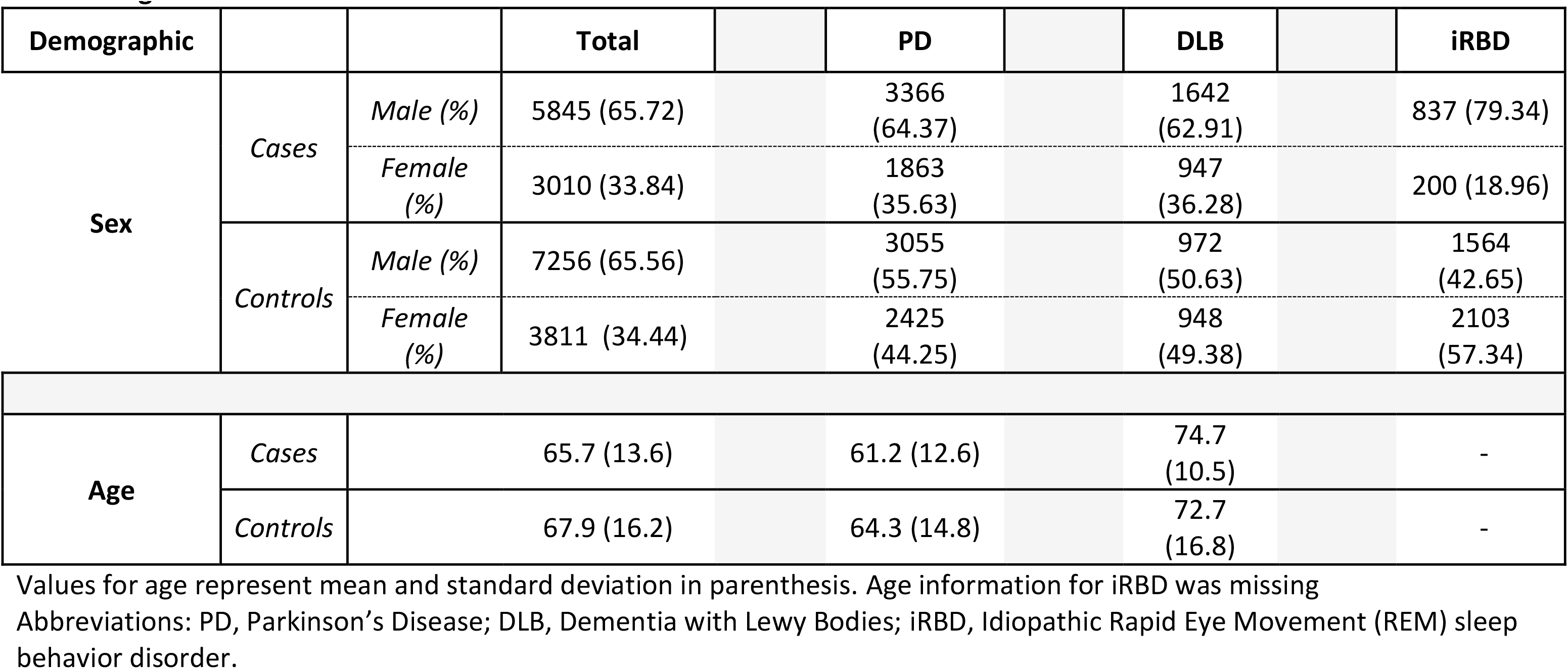
Age and sex of each cohort in cases and controls.

**Table 2:** *APOE* Haplotype Frequencies of each cohort in cases and controls.

| Group | Haplotype | Controls | Cases | Total |
| --- | --- | --- | --- | --- |
| DLB | e2e2 | 4 (.21) | 5 (.19) | 9 |
|  | e2e3 | 171 (8.91) | 189 (4.17) | 360 |
|  | e2e4 | 17 (.89) | 83 (3.18) | 100 |
|  | e3e3 | 1273 (66.30) | 1203 (46.09) | 2476 |
|  | e3e4 | 419 (21.82) | 916 (35.10) | 1335 |
|  | e4e4 | 36 (1.88) | 214 (8.20) | 250 |
|  | **Total** | **1920 (42.38)** | **2610 (57.62)** | **4530** |
| iRBD | e2e2 | 22 (.61) | 5 (.48) | 27 |
|  | e2e3 | 444 (12.27) | 109 (10.4) | 553 |
|  | e2e4 | 82 (2.27) | 15 (1.43) | 97 |
|  | e3e3 | 2241 (61.92) | 671 (64.03) | 2912 |
|  | e3e4 | 752 (20.78) | 236 (22.52) | 988 |
|  | e4e4 | 78 (2.16) | 12 (1.15) | 90 |
|  | **Total** | **3619 (77.54)** | **1048 (22.46)** | **4667** |
| PD | e2e2 | 41 (.75) | 25 (.48) | 66 |
|  | e2e3 | 725 (13.23) | 656 (12.55) | 1381 |
|  | e2e4 | 106 (1.93) | 75 (1.43) | 181 |
|  | e3e3 | 3599 (65.68) | 3498 (66.90) | 7097 |
|  | e3e4 | 954 (17.41) | 932 (17.82) | 1886 |
|  | e4e4 | 55 (1.00) | 43 (.82) | 98 |
|  | **Total** | **5480 (51.17)** | **5229 (48.83)** | **10709** |

Values represent frequency and percentage.
Abbreviations: PD, Parkinson's Disease; DLB, Dementia with Lewy Bodies; iRBD, Idiopathic Rapid Eye Movement (REM) sleep behavior disorder.

To further explore all potential genetic interactions between *APOE* and *SNCA*, we expanded the range to include all SNPs in *APOE* and *SNCA* plus 200kb outside the region ends on both genes across all cohorts. We pruned SNPs that were in LD with an r^2^ of 0.5 with 316 SNPs remaining in PD; 421 SNPs remaining in DLB; and 198 SNPs remaining in iRBD. We applied PLINKs regression-based approach to model and test SNP x SNP interactions. After correction for multiple comparisons with a Bonferroni correction, we did not identify any interactions associated with any of the alpha-synucleinopathies (Table 3).

**Table 3:** Top Hits of *SNCA* and *APOE* Haplotype Interaction Regression Results.

| Disease | Gene | Chr | Location | Ref Allele | Alt Allele | rs |  | Estimate | Standard Error | z-score | Pr(>\|z\|) |
| --- | --- | --- | --- | --- | --- | --- | --- | --- | --- | --- | --- |
| iRBD | SNCA | 4 | 90471245 | T | A | rs2870004 | rs2870004 x APOE Haplotype Interaction | -0.06359 | 0.06976 | -0.912 | 0.362 |
|  |  | 4 | 90626111 | G | A | rs356182 | rs356182 x APOE Haplotype Interaction | 0.03883 | 0.0629 | 0.617 | 0.537021 |
|  |  | 4 | 90756550 | C | G | rs7681440 | rs7681440 - APOE Haplotype Interaction | 0.04838 | 0.05954 | 0.812 | 0.416257 |
| PD | SNCA | 4 | 90471245 | T | A | rs2870004 | rs2870004 x APOE Haplotype Interaction | 0.039526 | 0.038801 | 1.019 | 0.30835 |
|  |  | 4 | 90626111 | G | A | rs356182 | rs356182 x APOE Haplotype Interaction | 0.027522 | 0.031979 | 0.861 | 0.389447 |
|  |  | 4 | 90756550 | C | G | rs7681440 | rs7681440 - APOE Haplotype Interaction | -0.016695 | 0.030556 | -0.546 | 0.584807 |
| DLB | SNCA | 4 | 89550094 | T | A | rs2870004 | rs2870004 x APOE Haplotype Interaction | -0.139925 | 0.118107 | -1.185 | 0.2361 |
|  |  | 4 | 89704960 | G | A | rs356182 | rs356182 x APOE Haplotype Interaction | 0.152286 | 0.110879 | 1.373 | 0.17 |
|  |  | 4 | 89835399 | C | G | rs7681440 | rs7681440 - APOE Haplotype Interaction | 0.070889 | 0.100602 | 0.705 | 0.481 |
| Abbreviations: PD, Parkinson’s Disease; DLB, Dementia with Lewy Bodies; iRBD, Idiopathic Rapid Eye Movement (REM) sleep behavior disorder. |  |  |  |  |  |  |  |  |  |  |  |

Abbreviations: PD, Parkinson's Disease; DLB, Dementia with Lewy Bodies; iRBD, Idiopathic Rapid Eye Movement (REM) sleep behavior disorder.

Lastly, a model-based Multifactor Dimensionality Reduction method was implemented as a final screen strategy to detect any significant SNP x SNP and SNP x SNP x SNP interactions using the open-source MB-MDR software^21^. Here too, no significant interactions were found to be associated with PD, DLB or iRBD after correction for multiple testing using permutation testing across all three cohorts (Supplementary Tables S1-S3).

## 4. Discussion

Our results do not support a role for genetic interactions between *APOE* and *SNCA* across PD, DLB, and iRBD. The genetic variants that were tested influence the expression and function of these proteins, but it is unlikely that any interactions between them affect the risk of developing synucleinopathies. Although functional epistasis, in the form of biomolecular interaction, can determine biological pathways of disease progression, it may not always be detected through mathematical or statistical genetic interaction analyses. Furthermore, the genetic interactions identified in our study, which represent three synucleinopathies, do not support the pathological associations observed in model organisms intended to represent these disease populations.

Previous studies have shown that APOE ε4 is linked to DLB in both AD and non-AD cases ^22,23^. However, some studies have also shown that APO ε4 is only associated with DLB when there is a significant amount of co-existing Alzheimer’s pathology ^24-26^. This finding contradicts the idea that APO ε4 independently drives α-synuclein pathology.

This study has several limitations. The lack of pathological confirmation of Lewy Bodies and AD pathology in the cohorts does not allow for an analysis based on co-pathology. Such analysis would have been able to detect interactions that exist only in subpopulation of patients, for example those who have both alpha-synuclein and amyloid pathology. Another limitation is that the study scope was limited to *APOE* and *SNCA*. Recently, a stratified GWAS of DLB uncovered an association only between *GBA* rs2230288 and pathologically confirmed DLB without AD pathology, but not in mixed pathological cases ^26^. This same SNP has been identified as a significant risk variant in the most recent GWAS of DLB ^7^. It is possible that large scale GWAS would uncover associations that encompass a broad spectrum of disease (e.g. DLB with no AD pathology, DLB with mixed PD and AD pathology etc.), even though these associations may be driven by subsets of these genetic loci (e.g *GBA*). Furthermore, it is plausible that different disease subtypes consist of distinct genetic combinations and interactions that have not yet been identified at a population level due to the limited size of current sample cohorts. Further studies should include more samples and implement alternative statistical or interrogative methods to leverage the current data to its fullest potential despite its small sample size.

## Data Availability

The data that support the findings of this study are available from dbGaP. Parkinson's patients and controls (phs000918.v1.p1). DLB patients and controls are available from dbGaP (phs001963.v1.p1). iRBD patients and controls are available upon reasonable request from the corresponding author. Additional controls were also obtained from dbGaP; NGRC (phs000196.v2.p1) and NIND (phs000089). Code used for analysis can be found on GitHub (https://github.com/gan-orlab/APOE_SNCA)

## Acknowledgements

We would like to thank all the participants in the different cohorts. We would also like to thank all members of the International Parkinson Disease Genomics Consortium (IPDGC). See for a complete overview of members, acknowledgements, and funding http://pdgenetics.org/partners.

## Funding

This work was financially supported by the Michael J. Fox Foundation and the Canadian Consortium on Neurodegeneration in Aging (CCNA). GAR holds a Canada Research Chair in Genetics of the Nervous System and the Wilder Penfield Chair in Neurosciences. EAF is supported by a Foundation Grant from the Canadian Institutes of Health Research (FDN grant – 154301). ZGO is supported by the Fonds de recherche du Québec - Santé (FRQS) Chercheurs-boursiers award and is a William Dawson Scholar.

## Competing Interests

Prabhjyot Saini - Nothing to declare

Eric Yu - Nothing to declare

Mehrdad A. Estiar - Nothing to declare

Lynne Krohn - Nothing to declare

Kheireddin Mufti - Nothing to declare

Uladzislau Rudakou - Nothing to declare

Jennifer A. Ruskey - Nothing to declare

Farnaz Asayesh - Nothing to declare Sandra B. Laurent - Nothing to declare Dan Spiegelman - Nothing to declare

Jean-François Trempe - Nothing to declare

Timothy G. Quinnell - Nothing to declare

Nicholas Oscroft - Nothing to declare

Isabelle Arnulf - I.A. was previously consultant for Idorsia pharma, and UCB Pharma.

Jacques Y. Montplaisir – Nothing to declare

Jean-François Gagnon - Jean-François Gagnon

Alex Desautels - Alex Desautels received operating grants from CHIR, AASM and research grants from Eisai, Takeda and Canopy Growth; honoraria from serving on the scientific advisory board of Eisai, Paladin Labs and UCB, as well as honoraria from speaking engagements from Eisai, Jazz Pharma and Paladin Labs. None of the financial disclosures is relevant to the submitted work.

Yves Dauvilliers - has served as a consultant or on advisory boards for Avadel Pharmaceuticals, Jazz Pharmaceuticals, UCB, Takeda Pharmaceutical Co., Theranexus, Harmony Biosciences, Bioprojet Pharma, and Idorsia

Gian Luigi Gigli - Nothing to declare

Mariarosaria Valente - Nothing to declare

Francesco Janes - Nothing to declare

Andrea Bernardini - Nothing to declare

Karel Sonka - Nothing to declare

David Kemlink – Nothing to declare

Wolfgang Oertel - Wolfgang H. Oertel has received speaker’s honoria on educational symposia sponsored by Abbvie, the International Movement Disorders Society and Stada Pharma. He acts as consultant for Lario Therapeutics and is a member of advisory boards with Intrabio and MODAG. He holds stock options with Intrabio not related to this manuscript and stock options with MODAG not related to this work. The institution of W.H.O., not W.H.O personally received/s scientific grants from the Stichting ParkinsonFonds The Netherlands, The ParkinsonFonds Germany related to this manuscript and scientific grants from the German Research Foundation, the Michael J Fox Foundation and the Rittal Foundation unrelated to the manuscript.

Annette Janzen -received grants from the ParkinsonFond Deutschland.

Giuseppe Plazzi – Has received consultancy fees for Bioprojet, Jazz, Takeda, Idorsia, Alkermes, and Centessa.

Elena Antelmi – Nothing to declare

Francesco Biscarini – Honorarium from BioProjet

Michela Figorilli – Nothing to declare

Monica Puligheddu – Nothing to declare

Brit Mollenhauer – Nothing to declare

Claudia Trenkwalder – Nothing to declare

Friederike Sixel-Döring – Nothing to declare

Valérie Cochen De Cock – Nothing to declare

Christelle Charley Monaca – Nothing to declare

Donald Grosset – Nothing to declare

Anna Heidbreder - Nothing to declare

Luigi Ferini-Strambi - Nothing to declare

Femke Dijkstra – Nothing to declare

Mineke Viaene – Nothing to declare

Beatriz Abril – Nothing to declare.

Bradley F. Boeve - Honorarium for SAB activities for the Tau Consortium - funded by the Rainwater Charitable Foundation; institutional research grant support for clinical trials from Alector, Transposon, Cognition Therapeutics, EIP Pharma; grant support from NIH, Lewy Body Dementia Association, American Brain Foundation, Mayo Clinic Dorothy and Harry T.

Mangurian Jr. Lewy Body Dementia Program, the Little Family Foundation, the Ted Turner and Family Foundation

Ronald B. Postuma - R.B.P. reports grants and personal fees from Fonds de la Recherche en Sante, grants from Canadian Institute of Health Research, The Michael J. Fox Foundation (MJFF), the Webster Foundation, Roche, and the National Institute of Health and personal fees from Takeda, Biogen, AbbVie, Curasen, Lilly, Novartis, Eisai, Paladin, Merck, Vaxxinity, Korro, Bristol Myers Squibb and the International Parkinson and Movement Disorders Society, outside the submitted work.

Guy A. Rouleau – Nothing to declare

Abubaker Ibrahim - Nothing to declare

Ambra Stefani -Nothing to declare

Birgit Högl – Nothing to declare

Michele T.M. Hu - Nothing to declare

Sonja W. Scholz - S.W.S. serves on the scientific advisory board of the Lewy Body Dementia Association, Mission MSA, and G-Can. S.W.S. receives research support from Cerevel Therapeutics.

Ziv Gan-Or received consultancy fees from Lysosomal Therapeutics Inc. (LTI), Idorsia, Prevail Therapeutics, Ono Therapeutics, Denali, Handl Therapeutics, Neuron23, Bial Biotech, Bial, UCB, Capsida, Vanqua bio, Congruence Therapeutics, Takeda, Jazz pharmaceuticals, Guidepoint, Lighthouse and Deerfield

## Notes

### Author Declarations

Informed consent and ethics approval was obtained from the appropriate institutional review boards at participating institutions as described in the original studies.

### Summary of Updates

Four authors were removed that were mistakenly added to the authorship list.

